# A first-in-human clinical study of an intranasal spray of a cocktail containing two synergetic antibodies neutralizes Omicron BA.4/5

**DOI:** 10.1101/2023.03.17.23287398

**Authors:** Xinghai Zhang, Feiyang Luo, Huajun Zhang, Hangtian Guo, Junhui Zhou, Tingting Li, Shaohong Chen, Shuyi Song, Meiying Shen, Yan Wu, Yan Gao, Xiaojian Han, Yingming Wang, Chao Hu, Xiaodong Zhao, Huilin Guo, Dazhi Zhang, Yuchi Lu, Wei Wang, Kai Wang, Ni Tang, Tengchuan Jin, Menglu Ding, Shuhui Luo, Cuicui Lin, Tingting Lu, Bingxia Lu, Yang Tian, Chengyong Yang, Guofeng Cheng, Haitao Yang, Aishun Jin, Xiaoyun Ji, Rui Gong, Sandra Chiu, Ailong Huang

## Abstract

Neutralizing monoclonal antibodies (NAbs) with prophylactic and therapeutic efficacy have demonstrated fundamental importance in the control of SARS-CoV-2 transmission. However, their wide application has been largely limited by high cost and inconvenience in administration. Here, we developed an intranasal spray containing two synergetic human NAbs that could broadly neutralize the emerging Omicron variants in vitro. A unique synergetic neutralizing mechanism was identified that the two NAbs bound to exclusive epitopes on the RBD and structurally compensate each other in blocking the Spike-ACE2 interaction. Importantly, when given at low dosages for three consecutive days through the intranasal mucosal route, this cocktail showed significant improvement in the emergency preventive and therapeutic effects in hamsters challenged with authentic Omicron BA.1. Further, we performed an investigator-initiated trail in healthy volunteers (ChiCTR2200066525) to study the safety and pharmacokinetics of the antibody cocktail administrated as nasal spray. The nasal spray is generally safe and well tolerated without treatment related severe abnormal effects. The antibody cocktail nasal spray demonstrated nasal concentrations higher than the IC_90_ of neutralization activity against Omicron BA.4/5 even at 24 hours post dosing. Furthermore, nasal samples from the study subjects demonstrated potent neutralization activity against Omicron BA.4/5 in an ex vivo pseudovirus neutralization assay. Together, we provide a novel approach for NAb regimens, a potentially highly effective product with broad applicable perspective in depressing the infection risk of new epidemic variant and ameliorating the heavy medical burden of hospital.

**One Sentence Summary:** An intranasal spray of two synergetic antibodies cocktail neutralizing Omicron BA.4/5 and an initial clinical evaluation in healthy volunteers.

## INTRODUCTION

Since the outburst of COVID-19 in December 2019, SARS-CoV-2 continues to evolve substantially, acquiring sets of mutations that enhance its transmissibility, infectivity and the ability to escape natural and acquired immunity (*1–3*). Contrast to other variants that preferentially engage in the lungs, Omicron alters the route of viral entry into host cells and is prompted to replicate in the upper airway (*4–6*). This causes more asymptomatic infections contributing to silent spread of the virus and poses substantial difficulties for effective prevention for the spread of infection (*7–9*). Omicron and its sub variants exhibit dramatic antigen shift, which have been shown to render the booster vaccination or recovery sera ineffective and cause breakthrough infections (*10–13*). Recent studies further hampered the active immune protection with evidences of inadequate protection against Omicron sublineages from vaccinated boosters based on Omicron BA.1 (*14–16*). Moreover, Paxlovid, a combination of two small molecule inhibitors recently received emergency authorization for patients at higher risk of critical illness, failed to prevent close contact infection in family members living with patients (*17–19*). Thus, alternative and supplemental prophylactic drugs are now urgently needed to prevent Omicron infection and subsequently block its transmission in the community (*20–22*).

Passive antibody administration based on neutralizing antibodies (NAbs) have demonstrated protective efficacy in susceptible individuals with moderated-to-severe immune compromise or vaccination contraindication, with promising potential in breaking the transmission chain (*23–25*). However, the high cost and the inconvenient intramuscular or intravenous way of administrations have drastically limited its application in broad population (*20, 26, 27*). To overcome these practical drawbacks, passive transfer inhibitors through the intranasal mucosal route may be promising approach to prevent the spread of Omicron (*28–31*). Antibodies delivery through an inhalant has been shown to facilitate early-stage contact with the pathogen in the respiratory tract (*32–34*). This may serve as an applicable platform enabling NAbs enrichment in the route of viral entry (e.g. nasal cavity and the upper airway), and overcoming the low and unsatisfying distribution of NAbs at the site of infection when administered routinely through a systemic route (*35–37*).

Here, we identified an intranasal applicable cocktail with broad neutralizing capability against Omicron and its sublineages, and determined its synergetic mechanism. We further investigated the prophylactic and treatment efficacy of this cocktail against Omicron BA.1 in a hamster model and evaluated its potential capability of the nasal spray convenient for self-administration to block the infection of BA.4/5 in a first-in-human trial.

## RESULTS

### Broad neutralizing ability of the cocktail containing 58G6 and 55A8

58G6 and 55A8 were two NAbs identified from COVID-19 convalescent patients at early 2019 (*31, 38–40*). 55A8 could bind to the spike (S) proteins of wide-type SARS-CoV-2 and its mutational variants, including Omicron BA.1, BA.2 and BA.4/5, as tested by enzyme-linked immunosorbent assay (ELISA) (fig. S1A and B). Biolayer interferometry (BLI) analysis revealed that 55A8 exhibited strong binding affinities to the S proteins of SARS-CoV-2, Delta and Omicron BA.5, and particularly for Omicron BA.1 and BA.2, at sub-picomolar level (<10^−12^ M) (fig. S1C). Further, we confirmed that cocktail of 55A8 and 58G6 could neutralize the pseudoviruses of SARS-CoV-2 variant strains (Fig. 1A, and fig. S2A and B). Interesting, the cocktail of 58G6 and 55A8 demonstrated obviously synergetic effects against pseudotyped SARS-CoV-2 variants as well as the authentic Omicron BA.1, with the half inhibition concentration exponentially lower than the currently approved NAbs for the emergence treatment of COVID-19 (Fig. 1A and B). Therefore, this cocktail consisted of potent neutralizers 58G6 and 55A8 with synergetic potency and breadth against the Omicron variants.

**Fig. 1.**
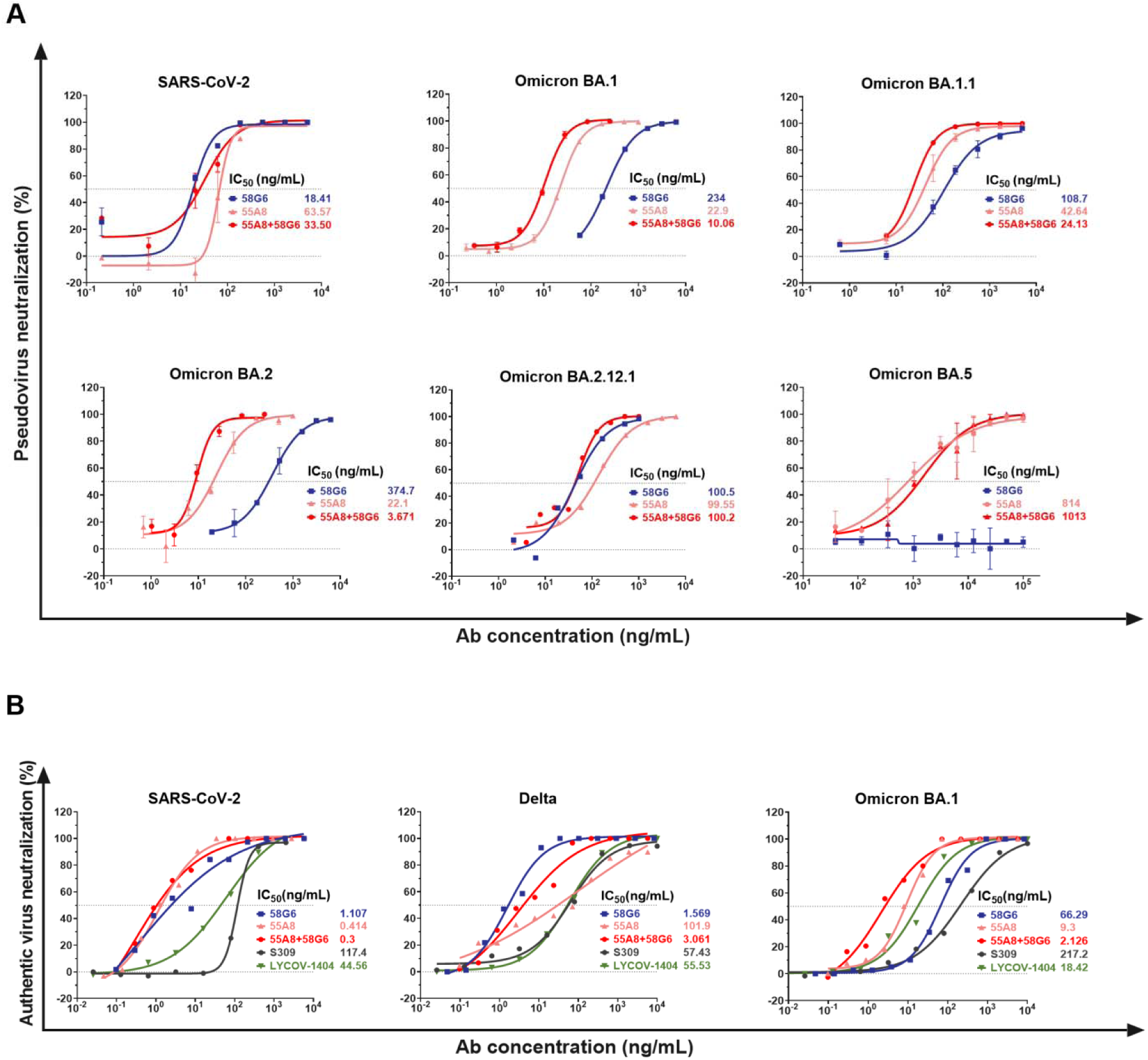
The cocktail of 58G6 and 55A8 broadly neutralizing Omicron emerging variants. (**A**) The neutralizing potencies of 58G6, 55A8 and cocktail of 58G6 and 55A8 against SARS-CoV-2, Omicron BA.1, BA.1.1, BA.2, BA.2.12.1 and BA.5 variants were measured with a pseudovirus neutralization assay. The dashed line indicates a 0% or 50% reduction in viral neutralization. Data for each NAb were obtained from a representative neutralization experiment of two replicates, presented as mean values ± SEM. (**B**) Neutralization against authentic SARS-CoV-2, Delta and Omicron BA.1 viruses.

### Synergetic neutralizing mechanism of this cocktail

To investigate the neutralizing mechanism of the 55A8, we first studied the single-particle cryo-electron microscopy (cryo-EM) structures of the antigen-binding fragments (Fabs) of 55A8 in complex with the prefusion Omicron BA.1 S trimer (fig. S3A). In all observed 55A8 Fabs-S complexes, the S trimer adopted a “1-up/2-down” or a “2-up/1-down” conformation (fig. S3A and B). Superimposition of the down RBD in the structures of the 55A8 Fab-S and ACE2-S complexes revealed that a 55A8 Fab binding with a down RBD could create a steric clash between ACE2 and the adjacent up RBDs, while no overlap between the 55A8 Fab and ACE2 on the same up RBD were observed (Fig. 2A).

**Fig. 2.**
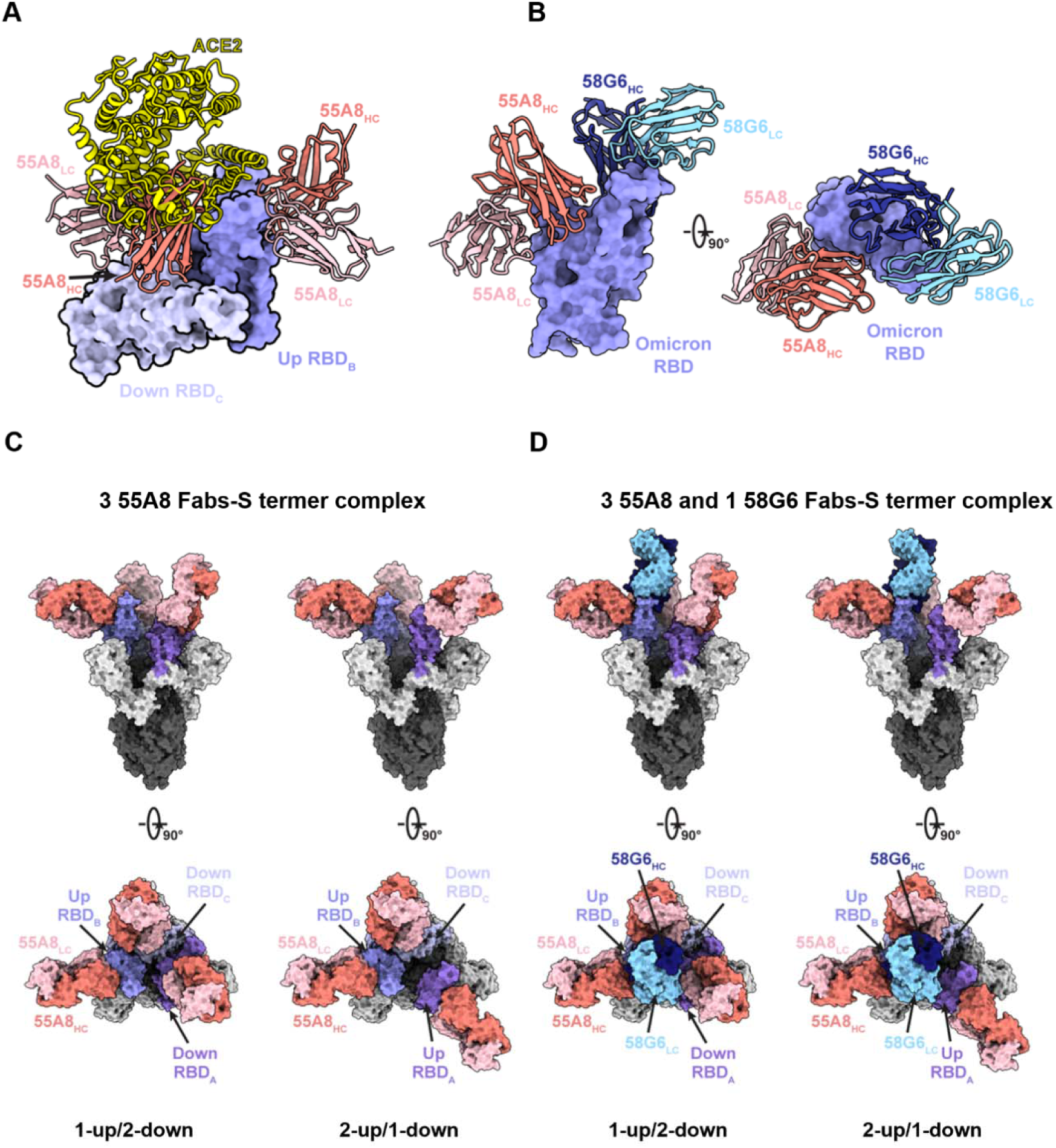
Synergetic neutralizing mechanism of the cocktail of 58G6 and 55A8. **(A**) Superposition of the locally refined Omicron RBD-ACE2 (PDB ID: 7WSA) model together with the locally refined Omicron RBD-55A8 Fab model. **(B**) Locally refined model of the 55A8 Fab and 58G6 Fab on the same up Omicron RBD. HC, heavy chain; LC, light chain. **(C**) 55A8 Fabs bind to Omicron S trimers in 2 states. Two perpendicular views of Omicron BA.1 S-55A8 complexes are shown as the surface. **(D**) 55A8 and 58G6 Fabs simultaneously bind to Omicron S trimers in 2 states. Two perpendicular views of Omicron S-55A8/58G6 complexes are shown as the surface. 55A8 heavy chain: salmon, 55A8 light chain: pink, 58G6 heavy chain: navy, 58G6 light chain: sky blue, three Omicron RBDs: different shades of purple.

Next, the cryo-EM structures of the Omicron BA.1 S trimer in complex with the 55A8 and 58G6 Fabs were determined and similar conformation of the BA.1 S trimer were observed when treated with the NAb-cocktail (fig. S3C). A refinement to an overall resolution of 3.3 Å showed that the majority of the selected particle images represented a 4-Fab-per-trimer complex, containing three 55A8 Fabs and one 58G6 (fig. S3D). Specifically, we found that 55A8 and 58G6 Fab simultaneously bound to a single up RBD, which exhibited no conformational changes comparing with the BA.1 S-55A8 Fab complex (Fig. 2B). The addition of 58G6 occupied the ACE2 binding site on the 55A8-bound RBD and further occluded the accessibility of the Omicron BA.1 S protein to ACE2 (Fig. 2C and D). This was further evidenced by the ACE2 competition assay, in which 55A8 showed no competition with ACE2 for binding to the Omicron BA.1 RBD, but partially competed with ACE2 for binding to the Omicron BA.1 S protein (fig. S4A). Also, we confirmed that 58G6 and 55A8 could simultaneously bind to the S proteins of SARS-CoV-2, Omicron BA.1 and Omicron BA.2 in a noncompetitive manner (fig. S4B). These findings suggested that the synergetic neutralization might be achieved through complementary steric occlusion of ACE2 by the pair of 55A8 and 58G6.

To determine the binding epitopes of these NAbs on Omicron BA.1 S proteins, we assessed the potential hydrogen bonds of 55A8 and 58G6 complementarity determining regions (CDRs) (fig. S5A to D). Specifically, half of the six (CDRs; CDRH3, CDRL1 and CDRL3) of the 55A8 Fab were found to directly participate in the interactions with the S^345-352^ and S^440-450^ regions (fig. S6A and B). Several potential hydrogen bonds, including R346, Y351, K440, S443, K444, V445 and N450, were identified at the interface of the 55A8 Fab and Omicron RBD (fig. S6C). The CDRs of 58G6 were shown to form unique interactions with the mutated amino acids N477, K478 and R493 within Omicron BA.1 RBM, explaining the sustained neutralizing capability of this RBM-targeted NAb against the Omicron BA.1 variants (fig. S6D).

### Intranasal delivery of this cocktail protect hamster from Omicron challenge

The protective efficacy of the cocktail of 55A8 and 58G6 against Omicron BA.1 infection were verified in a hamster model. On Day 0, hamsters received a single intranasal administration of 1500 μg 58G6, 500 μg 55A8 or combination of 300 μg 55A8 with 1000 μg 58G6 one hour prior to a challenge with 10^4^ plaque forming unit (PFU) of Omicron BA.1 through nasal drips (Fig. 3A). Post-infection, two additional NAb(s) treatments were given at day 1 and 2, and the animals were sacrificed one day later (Fig. 3A). RT–qPCR and plaque assays with harvested trachea and lungs tissues showed that viral RNA copies and infectious virus loads were significantly reduced (Fig. 3B and C). Importantly, the cocktail treatment resulted in robust viral clearance both in the upper respiratory tract (turbinate and trachea) and lower respiratory tract (lungs).

**Fig. 3.**
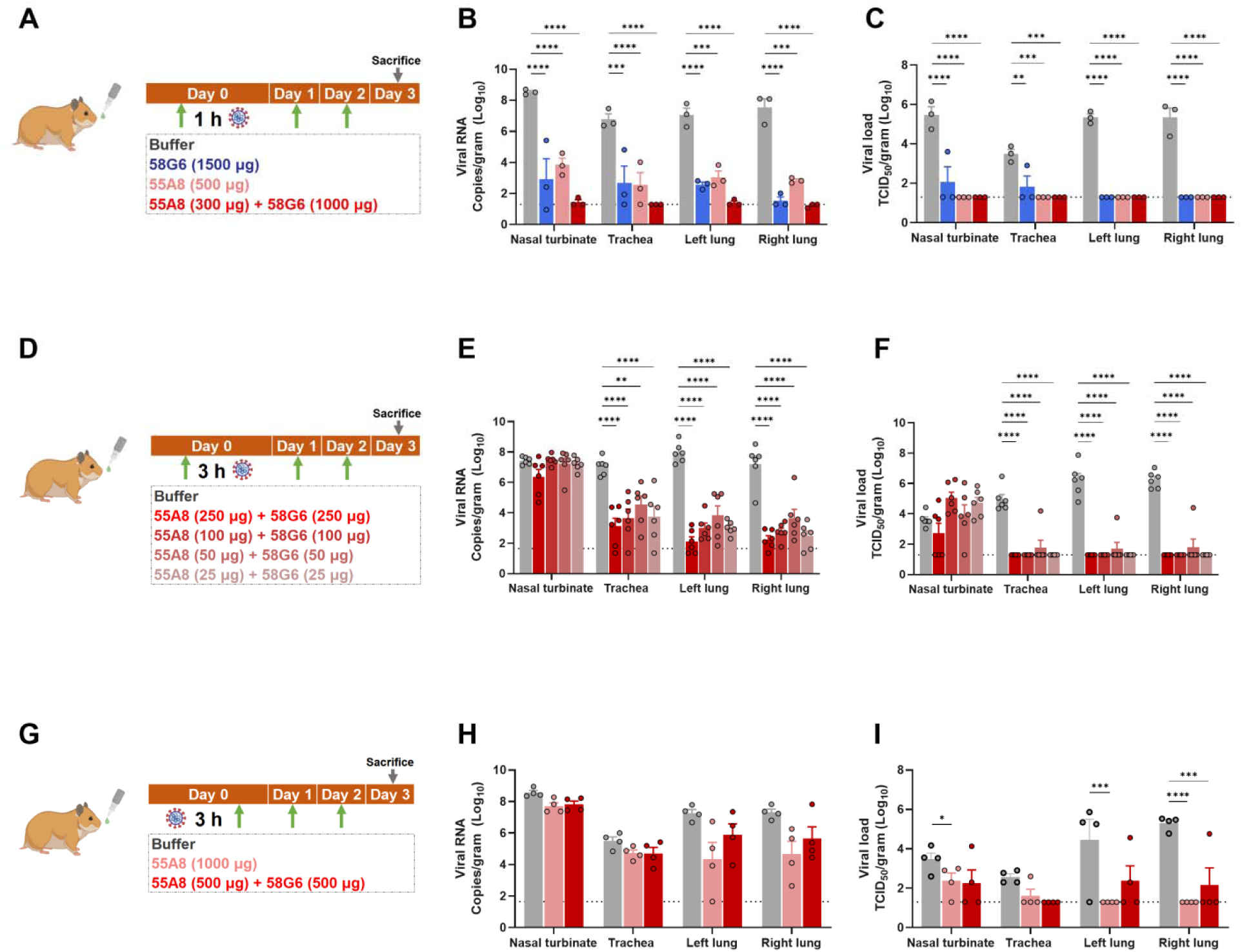
Intranasal delivery of cocktail of 55A8 and 58G6 protect hamster from Omicron challenge. Syrian golden hamsters challenged with 10^4^ PFU of Omicron were treated with 58G6, 55A8 or the two-antibody cocktail at 1 h or 3 hrs pre-infection, or 3 hrs post-infection, and two additional treatments at 24 and 48 h post-infection. The turbinates, trachea and lungs were harvested on day 3 post-treatment and analyzed by viral RNA and infectious virus (by qRT–PCR and PFU/g). (**A**), (**D**) and (**G**) Animal experimental scheme. (**B**), (**E**) and (**H**) Viral RNA (log_10_(RNA copies per g)) was detected in the respiratory tract of hamsters (**C**), (**F**) and (**I**) The infectious viruses (PFU) in the respiratory tract was measured with a viral plaque assay performed with Vero E6 cells.

Further, we investigated the protective efficacy of this cocktail with lower dosages and a prolonged pretreatment time (Fig. 3D). Significant decrease in the viral RNA copies were found in the trachea and lung tissues, and the infectious viral load was reduced substantially even with remarkably low dose of the cocktail treatment (25 μg 55A8 + 25 μg 58G6) (Fig. 3E and F). Encouraged by the observed protective effects associated with the cocktail spray, we analyzed the emergency treatment potential of 55A8 alone and the cocktail at 3 h post-infection (Fig. 3G). Expectedly, post-infection administration of 55A8 or the cocktail did not affect viral copies detected in the airway tissues (Fig. 3H). However, 55A8 profoundly impacted successful viral assembly, indicated by the significant reduction of virus load in both lungs (Fig. 3I). These data showed that the cocktail of 55A8 and 58G6 could confer protection even at markedly low doses and displayed a potential applicable value in the emergency treatment through the convenient administration with intranasal delivery.

### First-in-human trial of 55A8 and 58G6 antibody cocktail in healthy volunteers

A cocktail of 55A8 and 58G6 at 4:1 mass ratio was selected based on neutralization activity against Omicron BA.4/5 pseudovirus (Table S1). A formulation of 5 mg/mL total antibody concentration for the cocktail filled in a nasal spray device for self-administration was produced under Good Manufacturing Practice (GMP). This 4:1 combination of 55A8 and 58G6 at 5 mg/mL is called “A8G6” antibody cocktail. A8G6 cocktail had studied for preclinical toxicity in Rhesus monkeys under Good Laboratory Practice (GLP) and no safety concern was observed. A first-in-human randomized and placebo-controlled trial on the intranasal delivery of the antibody cocktail was conducted in 108 healthy volunteers (Fig. 4) to assess the safety/tolerability (primary objective) and pharmacokinetics (nasal and serum concentration over time, secondary objective) of A8G6 nasal spray. The baseline demographic information of the study subjects is listed in Table S2 and A8G6 versus placebo is 5 to 1 in all cohorts. The trial started with cohort 1, in which different doses (ranging from one dose to four doses per day) were given within one day (Fig. 4). In cohort 2, 3 and 4, four doses per day were given for 3, 7 and 14 consecutive days. Overall, A8G6 nasal spray is well tolerated with minimum treatment related adverse effects (Table S3 and S4) and the primary objective was met.

**Fig. 4.**
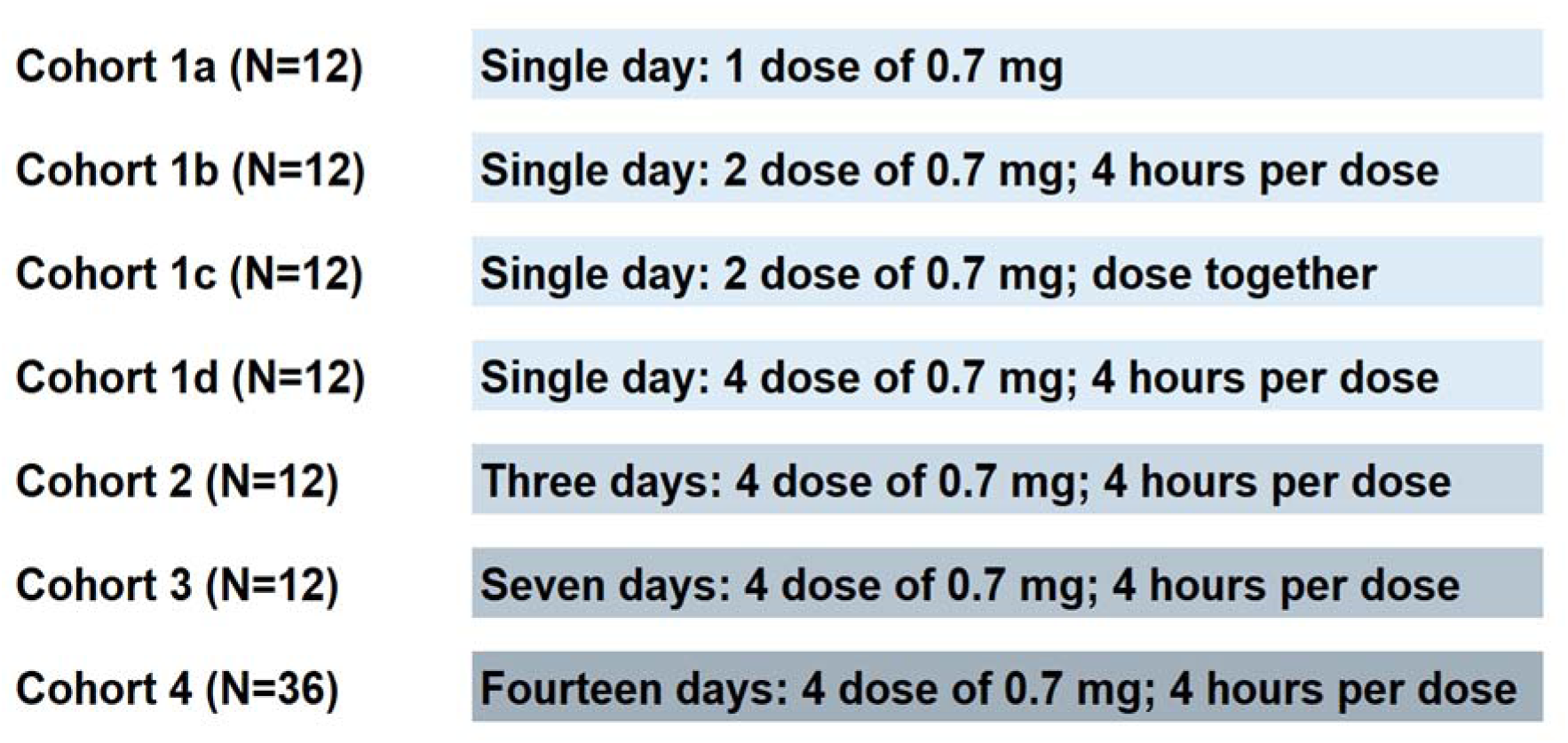
Trial design for first-in-human study of 55A8/58G6 (A8G6) nasal spray neutralization antibody cocktail. A total of 108 healthy volunteers were enrolled in 4 cohorts of study. A8G6: Placebo = 5:1 for each cohort. Cohort 1 has 4 sub-cohorts, which focused on the study of different doses (1, 2, 4 doses) within one day of dosing. Cohort 2 to 4 focused on 4 doses per day over 3, 7 and 14 days, respectively.

In second objective of the trial, the pharmacokinetics of A8G6 nasal spray was studied to optimize the dosing regimen. The target concentration is defined as nasal concentration above BA.4/5 pseudovirus neutralization IC_90_, which is about 5000 ng/mL. The results showed that after a single dose nasal administration, nasal concentration of A8G6 can maintain above BA.4/5 neutralization IC_90_ for 8 hours in > 90% subjects and dropped to below BA.5 IC_90_ at 24 hours post single dose for >50% of subjects (Fig. 5A). This suggests that multiple doses are required to maintain nasal concentration above BA.4/5 IC_90_ throughout the entire day. Similarly, two doses of nasal spray separated by 4 hours can maintain nasal concentration above IC_90_ 8 hours post 2^nd^ dose but cannot provide 24 hours coverage (Fig. 5B). When dosed at 2*0.7 mg per dose, nasal concentration of A8G6 was similar to 0.7 mg per dose (Fig. 5A and C). Four doses of A8G6 at 4 hours per dose, however, can achieve nasal concentration above BA.4/5 IC_90_ at 24 hours post 1^st^ dose or until the 1^st^ dose of next day (Fig. 5D). Collectively, cohort 1 data suggest that higher dose than 0.7 mg per dose does not result in higher nasal A8G6 concentration; repeat dosing during the day is required to maintain nasal concentration above the target concentration of BA.4/5 IC_90_.

**Fig. 5.**
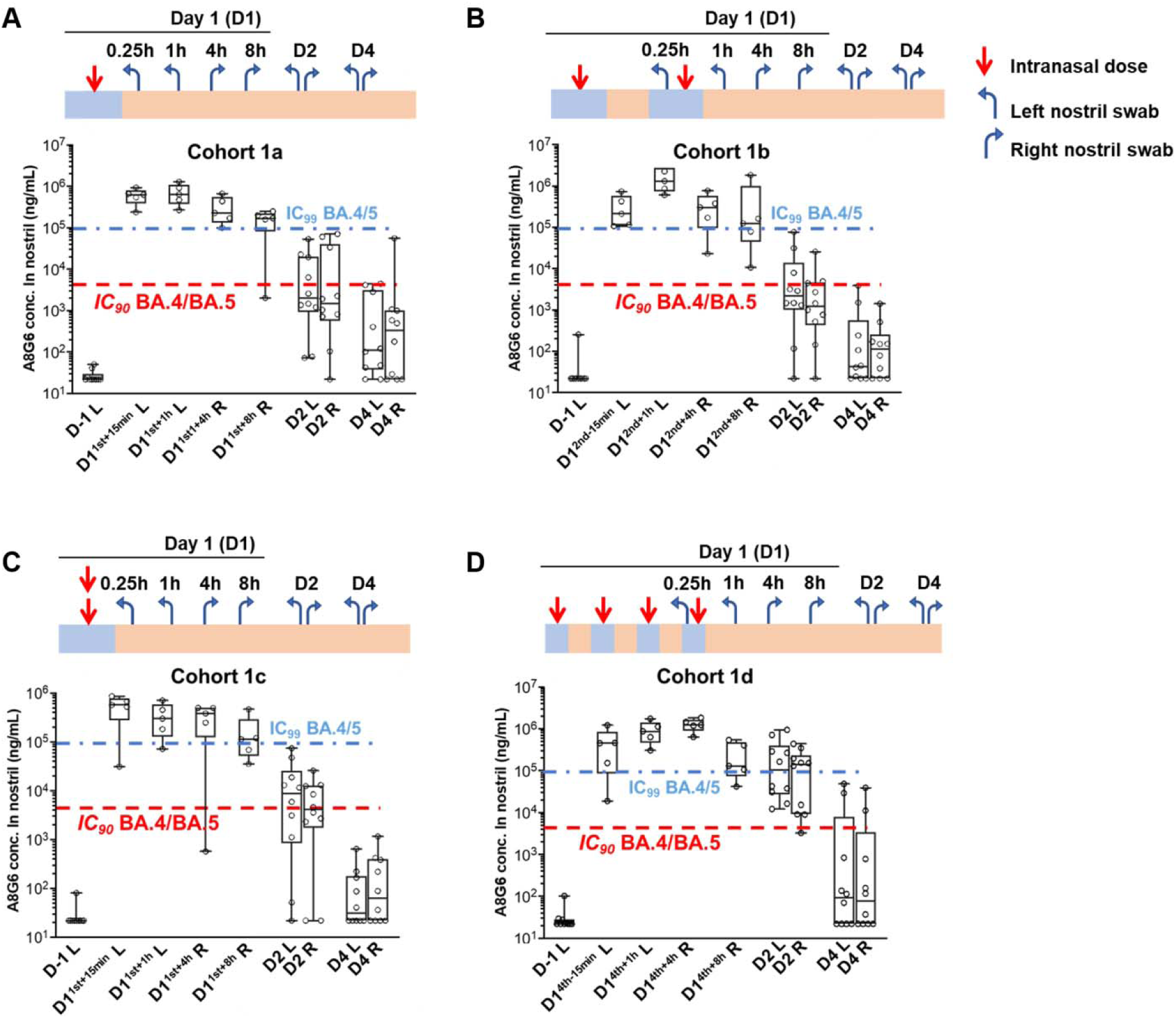
Cohort 1 clinical study of 55A8/58G6 (A8G6) cocktail showed nasal NAbs concentration above the IC_90_ neutralization activity on Omicron BA.4/5. concentration above the IC_90_ neutralization activity on Omicron BA.4/5. In each sub-cohort (1a, 1b, 1c, and 1d), 12 healthy volunteers were given different doses of cocktail NAbs (each dose is 560 μg of 55A8 plus 140 μg of 58G6, total 0.7 mg per dose) to both left and right nostril. A8G6 nasal spray: placebo nasal spray = 10:2. After A8G6 nasal spray, 5 volunteers took nasal samples at 0.25 hour post last dose at the left nostril, and at 1 hour post last dose at the right nostril; the other 5 took nasal samples at 4 hours post last dose at the left nostril and at 8 hours at the right nostril; 24 hours and 72 hours after 1st dose, all 10 volunteers took the nasal swab at both left and right nostril. Nasal swab (containing ~50 μL of nasal surface mucus) was washed in 500 μL PBS buffer before testing for A8G6 concentration using ELISA assay. Nasal A8G6 concentration was predicted from A8G6 PBS wash concentration by multiple a dilution factor of 11. Omicron BA.4/5 pseudovirus neutralization IC_90_ and IC_99_ values were labeled as dashed line and dash/dot line, respectively. X-axis label legend of Cohort 1a: D-1 L is the day before dosing; D1^1st+15min^ L is day 1 15 minutes after 1^st^ dose left nostril nasal swab; D1^1st+1h^ L is day 1 1 hour after 1^st^ dose left nostril nasal swab; D1^1st+4h^ R is day 1 4 hours after 1^st^ dose right nostril nasal swab; D1^1st+8h^ R is day 1 8 hours after 1^st^ dose right nostril nasal swab; D2 L is 24 hours after day 1 1^st^ dose left nostril swab; D2 R is 24 hours after day 1 1^st^ dose right nostril swab; D4 L is 72 hours after day 1 1^st^ dose left nostril swab; D4 R is 72 hours after day 1 1^st^ dose right nostril swab. X-axis label nomenclature of other Cohorts is similar to cohort1a.

Since each dose of 0.7 mg can maintain 8 hours coverage above BA.4/5 IC_90_, the dosing frequency could be changed to 3 doses per day at 8 hours per dose. When dosing for multiple consecutive days (3, 7 and 14 days), the nasal concentration at the end of the dosing was similar to day 1, suggesting minimum accumulation of A8G6 in nasal cavity over multiple days of dosing (Fig. 6A to C). To study the amount of A8G6 cocktail entering systemic blood circulation, we developed an ELISA assay that can specifically detect 58G6 in human serum using anti-58G6 specific antibodies. The results showed that for trail subjects in repeat dosing cohort 3 and 4, the serum 58G6 concentrations are below the detection limit (0.5 ng/mL) at the beginning and at the end of dosing period. This suggested that A8G6 has minimum penetration in the systemic blood circulation, which is consistent with the fact that antibody is a large molecule that has limited permeability through the nasal mucosa. Further, we studied the pharmacodynamics of A8G6 cocktail nasal spray by testing the nasal samples from the study subjects for ex vivo neutralization activity against Omicron BA.4/5 (Fig. 7). The results confirmed more than 90% BA.4/5 neutralization activity in all the samples. This human nasal pharmacokinetics/pharmacodynamics (PK/PD) data provided a good basis for potential clinical efficacy of this nasal spray for preventing Omicron BA.5 infections.

**Fig. 6.**
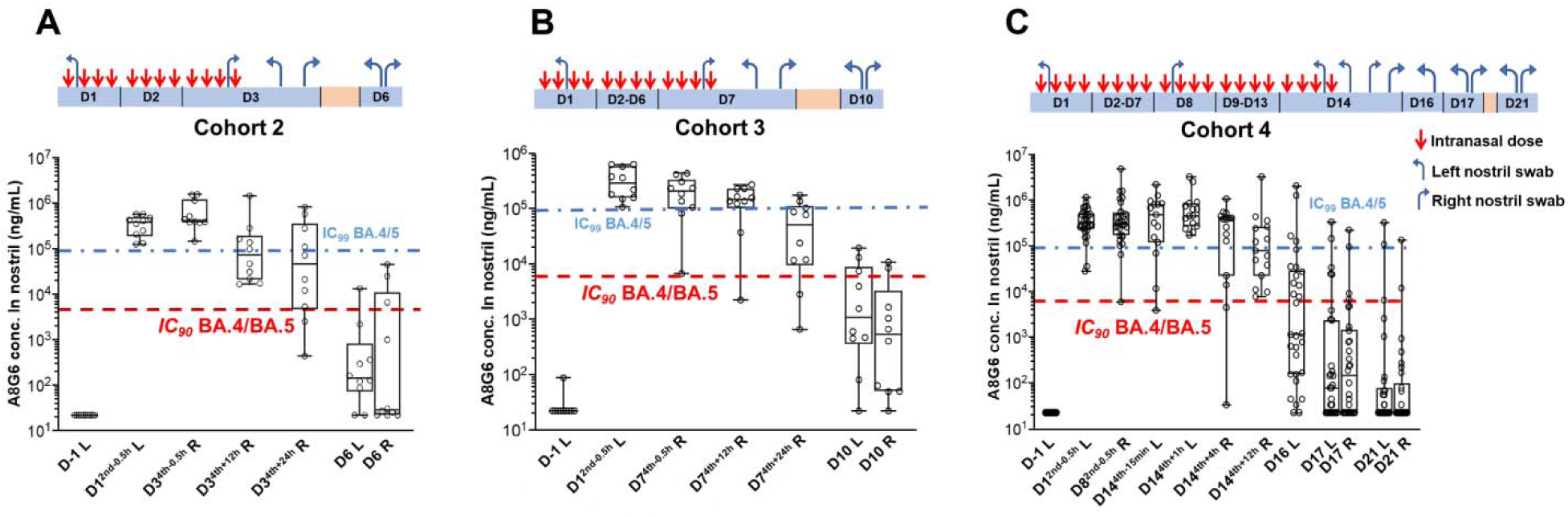
Cohort 2-4 clinical study of 55A8/58G6 (A8G6) cocktail showed nasal NAbs concentration above the IC_90_ neutralization activity on Omicron BA.4/5. In cohort 2-4, healthy volunteers were given 4 doses of cocktail NAbs (each dose is 560 μg of 55A8 plus 140 μg of 58G6, total 0.7 mg per dose) per day over 3, 7 and 14 days, respectively. A8G6 nasal spray: placebo nasal spray = 10: 2. After A8G6 nasal spray, 5 volunteers took nasal samples in alternate nostrils at each time point during dosing period, for a total of 4 time points that account for total of 10 volunteers. During the follow up period, all 10 volunteers took the nasal swab at both left and right nostril. Nasal swab (containing ~50 μL of nasal surface mucus) was washed in 500 μL PBS buffer before testing for A8G6 concentration using ELISA assay. Nasal A8G6 concentration was predicted from A8G6 PBS wash concentration by multiple a dilution factor of 11. Omicron BA.4/5 pseudovirus neutralization IC_90_ and IC_99_ values were labeled as dashed line and dash/dot line, respectively. X-axis label nomenclature is similar to Fig. 5.

**Fig. 7.**
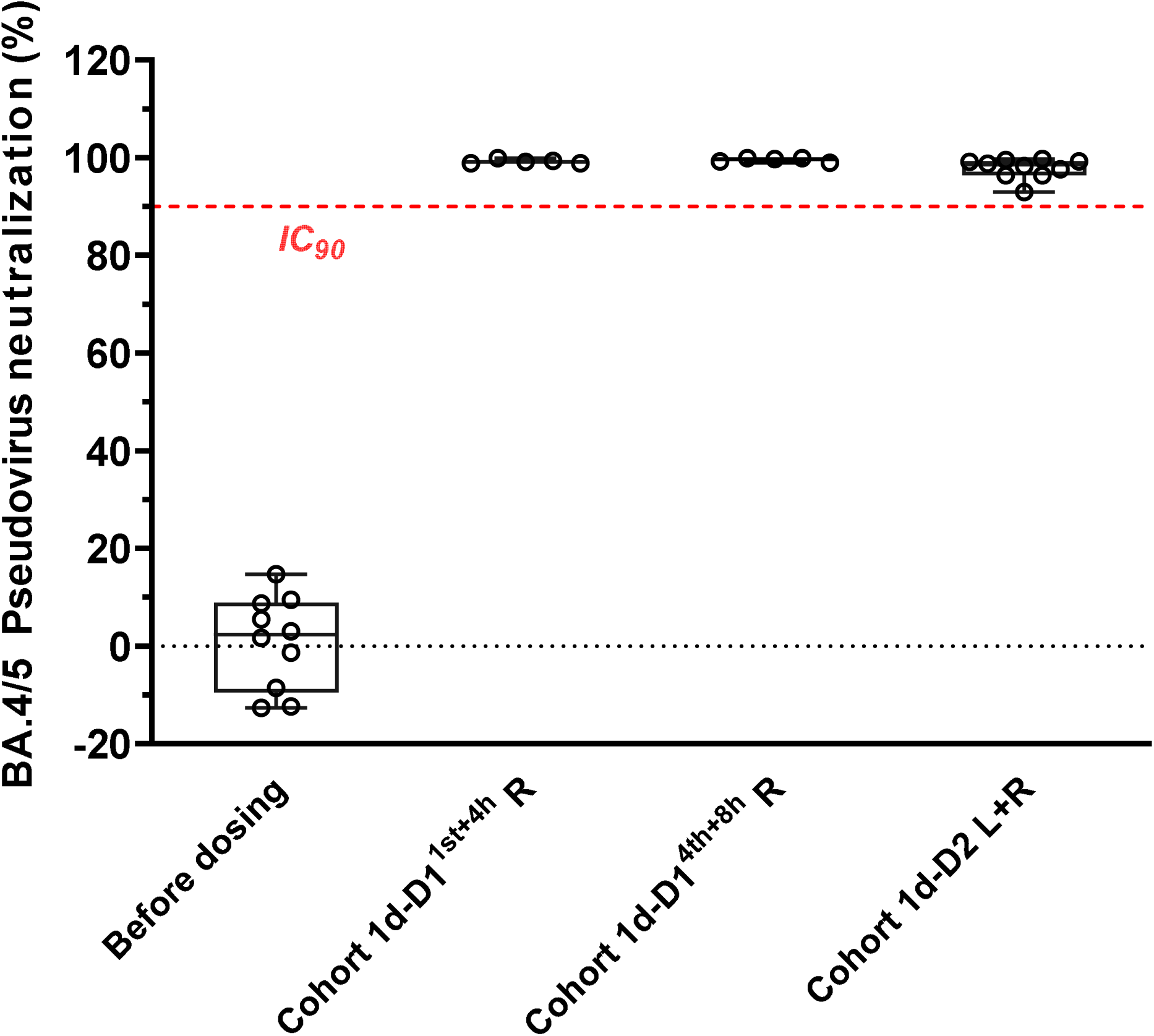
Selected nasal swab samples from cohort 1a and 1d showed > 90% neutralization activity on Omicron BA.4/5. Selected nasal samples from cohort 1a and cohort 1d were checked for BA.4/5 neutralization activities using a pseudovirus neutralization assay. Nasal samples were first diluted 11-fold in PBS then diluted 10-fold in cell culture media to measure their neutralization activities; the final neutralization activities in the original nasal samples were calculated by fitting a standard dose response curve.

## DISCUSSION

Given the identified rapid loss of protection by vaccination against SARS-CoV-2, the on-time NAb injection has been shown with timely prophylactic efficacy but has limited range of application, due to the high cost and inconvenience in administration (*20–22*). In this study, we identified two synergetic NAbs that could broadly neutralize the emerging Omicron variants. Synergetic neutralizing mechanism was found to largely rely on a unique complimentary addition of an RBM-targeted NAb (58G6) to a non-RBM NAb 55A8 to completely occlude ACE2 accessibility, with potential mutual inhibitions for the occurrence of escape mutations often seen with long-term application (*41–43*). This might greatly support the current lack of single NAb in face of Omicron and its sub variants that dramatically escape the majority of existing SARS-CoV-2 NAbs, and put the cocktail at the center stage for the development of clinically effective prophylactic regiments against the Omicron pandemic (*44*).

Together with the global attempt to provide affordable and accessible prophylactic drugs, we have advanced 55A8/58G6 (A8G6) cocktail into animal and human studies and presented the initial investigation by nasal spraying the NAb cocktail for the possibility of interrupting Omicron transmission in community (*20*). As the upper air tract has been shown to be favored by Omicron, several studies have proved evidences that intranasal pretreatment with small molecule inhibitor, anti-sense oligonucleotide (ASO) targeting SARS-CoV-2 RNA genome, anti-ACE2 mAb or mini-proteins mimicking hACE2 could effectively reduce respiratory virus replication and prevent SARS-CoV-2 infection (*28, 30, 32, 33, 45, 46*). It is evident that our A8G6 nasal spray antibody works uniquely differently from those nasal delivery molecules, by well-characterized mechanism of action and specificity, favorable NAb drug properties, or low safety risk with its local exposure. We further proved that the cocktail we identified could confer protective efficacy even at a markedly low dose of 50 μg against authentic Omicron BA.1 challenge in the hamster model, suggesting an economic outlook for the medical cost with its potential wide range applications. Together, all these features of A8G6 nasal spray provide strong foundation for fast development into clinical studies in human, while to the best of our knowledge, those other intranasal delivery molecules have not yet advanced into human studies or the clinical data have yet ready to be disclosed. This will make our study the first published detailed PK/PD data on a nasal spray of neutralizing antibodies for the potential prevention of Omicron, and the product based on this cocktail are currently undergone further clinical development to test for its applicable value in the prevention and emergency treatment against the Omicron pandemic (*44*).

The major challenge that arises with mucosal drug delivery comes from the rapid physical clearance by the mucociliary system (*47*). In response, repeated administrations of our NAbs and cocktail have shown with effective long-term protection, which may further reduce the potential risk of entrapping the virus in the mucosa by virus-antibody complexes. This calls for excellent biocompatibility with mAb products subjected for repetitive treatment. Hence, we chose patient-derived NAbs with low immunogenicity over other designed proteins or NAbs from other species, In addition, we utilized the IgG subtype of NAb to best restrict potential undesired immune responses. Although dimerized IgA are released into the upper airways and may represent elevated protective capacity and optimal on-site pharmacodynamics, IgG exhibit operational maturation in large-scale preparation with stable quality control between batches and are better choice for the potential wide range application of the NAb-based nasal spray in purpose of actual clinical use (*32*). Also, the fact that IgG is released to the lower lung through passive transudation may better facilitate the emergency treatment effect of our cocktail. Finally, the absence of immune response in the first-in-human trial of the cocktail NAb is encouraging for the concept of using repetitive intranasal protective measurements in the long combat with continuous emerging SARS-CoV-2 mutational variants.

There are a few limitations with our study. First, due to the lag in the availability of authentic Omicron sublineages variants, in vitro neutralization experiments were performed against wild-type SARS-CoV-2 and only two mutants, Delta and Omicron BA.1. Future experiments with updated Omicron sublineages are needed to determine the antiviral activity of our cocktail against the full landscape of the Omicron clade. Second, the physiological disparities of respiratory systems between human and rodent models raise questions for the observed preventative effects. Excitingly, as projected based on the faster respiration rate of rodents, cocktail 55A8/58G6 exhibited more persistent PK in human nasal cavity (Fig. 5A to D) than in rodents. Certainly, thorough efficacy study in human trials is warranted to validate the clinical efficacy of this cocktail.

In conclusion, we present a potent NAb cocktail against Omicron variants with high prophylactic efficacy at low dosage, which can be dosed in the self-administrable nasal spray format. The associated low cost and the needle-free convenience make it acceptable for potential large-scale application in general population. Our product represents promising passive NAbs interventions that may effectively aid the current prophylactic vaccines to mitigate the current SARS-CoV-2 transmission and its probable resurgence.

## MATERIALS AND METHODS

### Study design

The ultimate objective of this study was to collect the PK/PD information of an intranasal spray of a human antibody cocktail A8G6 data in a first-in-human trail. At the outset, human IgG antibodies with low immunogenicity were applied by means of intranasal spray and tested for the efficiency of blocking SARS-CoV-2 infection. Specifically, two nAbs (55A8 and 58G6) were selected from our screening library established before the dominance of Omicron BA.5, with outstanding neutralizing potency and broad neutralizing ability characterized by BLI affinity experiments and a standard pseudovirus and authentic virus neutralization assays. The specific neutralizing mechanism of 55A8 were further assessed by cryoEM. The observed synergetic neutralizing mechanism of 55A8 and 58G6 was addressed by structure analysis and confirmed by BLI competition assays of inhibiting RBD-ACE2 and Spike-ACE2 binding.

Next, we performed animal studies to determine whether low concentration of intranasal delivered 55A8 and 58G6 could provide protective efficacy pre- and post-exposure of Omicron BA.1 infection. Female hamsters (age of five to six weeks) were randomly assigned to groups. Animal researchers were not blinded to the study groups or during the assessment of the outcomes. No data points were omitted for the analysis. Sample sizes were determined based on previous experience. All the animal studies were reviewed and approved by the Institutional Animal Care and Use Committee of the Institute of Wuhan Institute of Virology, Chinese Academy of Sciences, and performed in an ABSL-3 facility (WIVA45202104).

This first-in-human trial was a randomized, double-blind, placebo-controlled study. A total of 108 healthy volunteers were enrolled into the study, each provided written informed consent in according with institutional guidelines. The trial is registered at Chinese Clinical Trial Registry with registration # ChiCTR2200066525. The trial was conducted in accordance with the Second Affiliated Hospital of Chongqing Medical University (Chongqing, China) Institutional Ethics Review Board (Study # AY-62-8001). The ethical approval of this trial complies with the requirements of the Good Clinical Practice, the Declaration of Helsinki of the World Medical Association, International Ethical Guidelines on Biomedical Research Involving Human Subjects of the Council for International Organizations of Medical Sciences, and relevant domestic laws and regulations. The sample size was determined based on precise assess both the primary and secondary objectives. The primary objective of this clinical study was to evaluate the safety and tolerance of A8G6 nasal spray under continuous repetitive use. To this end, we recorded the number of adverse events. The second objective was to evaluate the PK of A8G6 nasal spray to optimize the dosing regimen.

For PK analysis, we collected the blood samples at 1 and 3 days after final dose in cohort 1-3, and 3 and 7 days after final dose in cohort 4. Nasal swab was collected (LJ) at 0.25 and 4 hours after the final dose from left nostril, 4 and 8 hours from right nostril, and day 2 and day 4 from both nostrils in cohort 1a and 1c (Fig. 5A and 5C); (LJ) at 0.25 hours before and 4 hours after the final dose from left nostril, 4 and 8 hours after the final dose from right nostril, and day 2 and day 4 from both nostrils in cohort 1b and 1d (Fig. 5B and 5D); (LJ) at 0.5 hours before 2^nd^ dose on day 1 and 12 hours after last dose from left nostril, 0.5 hours before and 24 hours after last dose from right nostril, and day 6 from both nostrils in cohort 2 (Fig. 6A); (LJ) at 0.5 hours before 2^nd^ dose on day 1 and 12 hours after last dose from left nostril, 0.5 hours before and 24 hours after last dose from right nostril, and day 10 from both nostrils in cohort 3 (Fig. 6B); (LJ) at 0.5 hours before 2^nd^ dose on day 1, 15 mins before and 1 hours after last dose, and day 16 from left nostril, 0.5 hours before 2^nd^ dose on day 8, 4 and 12 hours after last dose from right nostril, and day 17 and 21 from both nostrils in cohort 4 (Fig. 6C). The concentrations of A8G6 from these samples were predicted by an ELISA assay containing a pair of specific anti-58G6 antibodies, and the corresponding neutralization activities of collected samples were measured using an Omicron BA.5 pseudovirus neutralization assay. An independent data safety monitoring committee performed trial oversight and made recommendations after review of safety reports between cohorts. Full details of the trial design, conduct, oversight and sample analysis and statistical analysis were provided in the protocol, which is available in the supplemental data.

### Statistical methods

Statistical analyses of the animal studies were performed using GraphPad Prism software v.9.2.0. Comparisons between two groups were performed using unpaired Student’s t tests. Comparisons among multiple groups were performed using one-way ANOVA followed by Tukey’s multiple comparison post hoc test. PLJ<LJ0.05 was considered significant (significance is denoted as follows: *PLJ<LJ0.05, **PLJ<LJ0.01, ***PLJ<LJ0.001, and ****PLJ<LJ0.0001).

## List of Supplementary Materials

### Materials and Methods

Figs. S1 to S6

Tables S1 to S5

A8G6 IIT Protocol

References (*48–60*)

## Data Availability

All data produced in the present study are available upon reasonable request to the authors

## Acknowledgments

We thank the Center for Animal Experiment and BSL-3 laboratory, Wuhan Institute of Virology, Chinese Academy of Sciences; Center for Biosafety Mega-Science, Chinese Academy of Sciences; and the National Virus Resource Center for resource support.

## Funding

The Natural Science Foundation of Hubei Province of China (2019CFA076)

The National Natural Science Foundation of China (32170949, 81871639, 92169109, 81871656 and 8181101099)

The National Science and Technology Major Project (2017ZX10202203)

The National Key Research and Development Program of China (2018YFA0507100 and 2016YFD0500300);

Guangzhou National Laboratory (SRPG22-015) Lingang Laboratory (LG202101-01-07)

Science and Technology Commission of Shanghai Municipality (YDZX20213100001556 and 20XD1422900).

The first-in-human Investigator Initiated Study was funded by the Emergency Project from the Science & Technology Commission of Chongqing (cstc2021jscx-fyzxX0001)

## Author contributions

A.H., S.C., A.J., H.Y., C.G. and C.Y. conceived and designed the project. For biological function analysis of the NAbs, F.L., T.L., M.S., X.H., Y.W. and C.H. screened and cloned the antibodies, and expressed and purified the antibodies; F.L., T.L. and W.W. were responsible for BLI assays for the binding ability, affinity, and the competition experiment of NAbs; F.L., T.L., S.S., K.W., N.T., M.D., and S. L. prepared various pseudovirus and conducted the pseudovirus neutralization assays. For the efficacy test of the NAbs *in vitro* and *in vivo*, X.Z., H.Z., J.Z., S.C., Y.W. and R.G. performed authentic SARS-CoV-2 neutralization assays and animal experiments. For structure analysis, H.G. and Y.L. cloned, expressed and purified Omicron BA.1 S proteins; H.G., Y.G., and Y.L. collected, processed the cryo-EM data, and built and refined the structure model; X.J., H.Y. and T.J. analyzed and discussed the cryo-EM data. A.H., M.D., S.L., C.L., T.L., B.L., Y.T., C.Y., and G.C. designed and supported the investigator-initiated trial. A. H., S.C., A.J., X.J., R.G., X.Z. and F.L., Y.T., G.C. wrote the manuscript. All authors revised and reviewed the final manuscript.

## Data and materials availability

The coordinates and cryo-EM map files for the 55A8-BA.1 S complexes and 55A8/58G6-BA.1 S complexes have been deposited in the Protein Data Bank (PDB) under accession number 7WWI, 7WWJ, 7WWK, 7XJ6, 7XJ8 and 7XJ9. All other data are available from the corresponding author upon reasonable request.

